# Genetic implication of prenatal GABAergic and cholinergic neuron development in susceptibility to schizophrenia

**DOI:** 10.1101/2023.12.14.23299948

**Authors:** Darren Cameron, Ngoc-Nga Vinh, Parinda Prapaiwongs, Elizabeth A. Perry, James T. R. Walters, Meng Li, Michael C. O’Donovan, Nicholas J. Bray

## Abstract

**Background:** The ganglionic eminences are fetal-specific structures that give rise to gamma-aminobutyric acid (GABA)- and acetylcholine- releasing neurons of the forebrain. Given evidence for GABAergic and cholinergic disturbances in schizophrenia, as well as an early neurodevelopmental component to the disorder, we tested the potential involvement of developing cells of the ganglionic eminences in mediating genetic risk for the condition.

**Study Design:** We combined data from a recent large-scale genome-wide association study of schizophrenia with single cell RNA sequencing data from the human ganglionic eminences to test enrichment of schizophrenia risk variation in genes with high expression specificity for particular developing cell populations within these structures. We additionally performed the single nuclei Assay for Transposase-Accessible Chromatin with Sequencing (snATAC-Seq) to map potential regulatory genomic regions operating in individual cell populations of the human ganglionic eminences, using these to additionally test for enrichment of schizophrenia common genetic variant liability and to functionally annotate non-coding variants associated with the disorder.

**Study Results:** Schizophrenia common variant liability was enriched in genes with high expression specificity for developing neuron populations that are predicted to form dopamine D1 and D2 receptor expressing GABAergic medium spiny neurons of the striatum, cortical somatostatin-positive GABAergic interneurons, calretinin-positive GABAergic neurons and cholinergic neurons. Consistent with these findings, schizophrenia genetic risk was also concentrated in predicted regulatory genomic sequence mapped in developing neuronal populations of the ganglionic eminences.

**Conclusions:** Our study provides evidence for a role of prenatal GABAergic and cholinergic neuron development in later susceptibility to schizophrenia.

## Introduction

The ganglionic eminences (GE) are transient fetal brain structures that are a major origin of gamma-aminobutyric acid (GABA)- and acetylcholine- releasing neurons of the forebrain. The GE can be divided into 3 regions (lateral, medial and caudal), with the lateral portion (LGE) generating GABAergic medium spiny neurons of the striatum, the medial portion (MGE) generating cortical and striatal GABAergic interneurons, and the caudal portion (CGE) generating distinct populations of GABAergic neurons of the cerebral cortex and limbic system ^1–3^. Cells originating from the MGE additionally form cholinergic interneurons of the striatum and basal forebrain cholinergic projection neurons, the latter modulating activity in the cerebral cortex ^4^.

Reductions in GABAergic neuron markers (e.g. GAD67, parvalbumin and somatostatin) within the prefrontal cortex are one of the most replicated post-mortem findings in schizophrenia ^5–8^. These findings appear to reflect altered functioning, rather than loss, of subpopulations of these neurons ^5,8^ and have been hypothesized to have a developmental basis ^9–11^. Disturbances in cholinergic neuron function have also been postulated to play a role in the pathophysiology of schizophrenia ^12–14^, with dysfunction in either system plausibly contributing to the diverse symptomatology of the disorder.

Schizophrenia is known to have a strong genetic component ^15^, involving numerous susceptibility variants that are common in the general population ^16,17^ as well as rarer variants that can confer a greater effect on risk ^18,19^. Tissues and cell types of etiological relevance to a disorder can be delineated by testing whether genes that are prominently expressed within them are enriched for genetic association with the condition ^20–22^. It is now possible to profile global gene expression in individual cell populations from human tissues through single cell or single nuclei RNA sequencing (scRNA-Seq / snRNA-Seq). By combining this methodology with large-scale genome-wide association study (GWAS) data for schizophrenia ^16^, we have previously found that schizophrenia common variant liability is enriched not only in genes with high expression specificity for developing glutamatergic neurons, but also in genes that are prominently expressed in developing neurons of the GE ^23^. Here, we build on that work through use of an independent scRNA-Seq dataset derived from nearly 3 times as many cells from the human GE ^24^, allowing greater definition of the developing neuron populations derived from these structures that are likely to be relevant to schizophrenia pathogenesis. Given that the common genetic variants most strongly associated with schizophrenia are located predominantly in non-coding genomic sequence ^16^, we additionally perform the single nuclei Assay for Transposase-Accessible Chromatin with Sequencing (snATAC-Seq) on human fetal tissue to provide the first maps of open chromatin (an index of regulatory genomic regions) in individual cell populations from all 3 primary regions of the GE. By testing for enrichment of schizophrenia single nucleotide polymorphism (SNP) heritability in identified open chromatin regions, we find complementary evidence for a role of GE-derived neuron development in susceptibility to schizophrenia, and we further use these annotations to functionally prioritize non-coding genetic variants associated with the disorder.

## Methods

### Single cell RNA-Seq data

scRNA-Seq data from the whole GE of 6 human fetuses aged 9 – 18 gestational weeks (generated by Shi et al, 2021) ^24^ were downloaded from the Gene Expression Omnibus (accession GSE135827) as a matrix of raw unique molecular identifier (UMI) counts. The associated metadata were obtained from Supplementary Table 2 from Shi et al ^24^. RNA sequence alignment and quality control procedures, including doublet removal and cell / gene thresholding, are described by Shi et al ^24^. Cells that were labelled by the original authors ^24^ as deriving from tissue outside of the GE (‘excitatory neurons’, ‘excitatory intermediate progenitor cells’ or ‘thalamic neurons’) were excluded, leaving 30,484 cells for our analysis.

### scRNA-seq data processing

Preliminary scRNAseq data processing was carried out in Seurat, version 4.3.0 ^25^ (Hao et al., 2021), as described previously ^23^, with dimensionality reduction and batch correction conducted across samples using FastMNN ^26^. Cells were clustered according to gene expression profile using shared nearest neighbor modularity optimization-based clustering ^25^ and visualized in 2 dimensions via Uniform Manifold Approximation Projection (UMAP). Following Shi and colleagues ^24^, we considered GE cell types at 2 levels of granularity. Cells were initially categorized in broad (‘level 1’) terms into either developing neurons of the MGE (MGE-N), developing neurons of the LGE (LGE-N), developing neurons of the CGE (CGE-N), progenitor cells, intermediate progenitor cells (IPC) or microglia based on the expression of known cell markers. We then further refined the progenitor cells, IPCs and MGE, LGE, CGE neurons by sub-clustering each cell type, yielding a total of 36 (‘level 2’) sub-populations.

### Testing enrichment of schizophrenia common variant liability in genes with high expression specificity for individual cell populations of the GE

We removed uninformative (low and / or sporadically expressed) genes ^27^ and, due to its linkage disequilibrium structure, genes in the major histocompatibility complex (MHC) region (hg38; chr6:28,510,120-33,480,577). A cell specificity score was then calculated for each gene in each cell type by dividing the expression of the gene in that cell type by its summed expression across all cell types (a procedure performed separately for level 1 and level 2 cell types) ^27^ (Supplementary Tables S1 and S2). Following others ^16,22^ and our previous analysis in fetal brain ^23^, we used MAGMA (Multi-marker Analysis of GenoMic Annotation) ^28^ and SLDSR (Stratified Linkage Disequilibrium Score Regression) ^21^ to test for enrichment of schizophrenia common variant liability within genes in the top specificity decile for each cell population. Schizophrenia common variant association statistics were taken from the latest GWAS meta-analysis of the Psychiatric Genomics Consortium ^16^ (downloaded from https://figshare.com/articles/dataset/scz2021/14672178), based on data from >70,000 people with schizophrenia and >240,000 control individuals. We used a similarly powered GWAS for human height (N ∼ 700,000 individuals) ^29^, downloaded from https://portals.broadinstitute.org/collaboration/giant/index.php/GIANT_consortium_data_fi les, as a comparison non-brain phenotype. The boundaries of each gene’s transcribed region were extended by 35kb upstream and 10kb downstream for MAGMA and 100kb upstream and 100kb downstream for SLDSR, according to current recommendations ^21,28^. We additionally down-sampled the number of cells assigned to each population or used a fixed number of genes (the top 1000 most specific) for each cell type to assess the robustness of our findings. For SLDSR, significance was determined by calculating a Z-score based on whether schizophrenia single nucleotide polymorphism (SNP) heritability was enriched within each cell-specific gene set after controlling for 53 baseline genomic annotations, including genic, enhancer and conserved regions (baseline model version 1.2) ^30^. We primarily report cell-specific enrichments where one-tailed *P* values exceed the Bonferroni threshold (*P* < 8.3 X 10^-3^ for level 1 tests and *P* < 1.4 X 10^-3^ for level 2 tests) in both MAGMA and SLDSR.

### Single nuclei open chromatin profiling of the ganglionic eminences

Single nuclei ATAC-Seq (snATAC-Seq) was performed on whole GE from 3 human fetuses (two of 16 and one of 17 gestational weeks). Tissue was acquired from the MRC-Wellcome Trust Human Developmental Biology Resource (HDBR) (http://www.hdbr.org/) as fresh whole brain in Hibernate-E media (Thermo Fisher Scientific). Samples were obtained through elective terminations of pregnancy, with consent from female donors, and were of normal karyotype. Ethical approval for the collection and distribution of fetal material for scientific research was granted to the HDBR by the London - Fulham Research Ethics Committee and North East - Newcastle & North Tyneside 1 Research Ethics Committee. The whole GE were dissected from each sample under a light microscope and dounce homogenized on ice. Large debris and unlysed cells were pelleted by centrifugation at 100 RCF for 5 mins at 4°C and the supernatant strained through a 10µm filter (Cambridge Bioscience). Nuclei were then pelleted by centrifugation of the strained supernatant at 350 RCF for 5 mins. Pellets were then resuspended in buffer containing 10mM Tris-HCl (pH 7.4), 10mM NaCl, 3mM MgCl2, 0.1% Tween-20 and 1% BSA and incubated on ice for 5 mins. Nuclei were then again pelleted, resuspended in 1X Nuclei Buffer (10X Genomics), examined under a light microscope to ensure integrity and counted using an Invitrogen Countess II automated cell counter. snATAC-Seq libraries were prepared from an estimated 8,000 nuclei from each sample using Chromium Next GEM Single Cell ATAC (v1.1) reagents (10X Genomics), following manufacturer instructions. Quality control of libraries was performed using the Agilent 5200 Fragment Analyzer before sequencing on an Illumina NovaSeq 6000 to a depth of at least 617 million read pairs per library.

### snATAC-seq data processing

Raw sequencing data were converted into FastQ files using bcl2fastq (Illumina, version 2.18). snATAC-Seq reads were aligned to the human reference genome (hg38) and fragment files generated for each sample using cellranger-count-atac (10X genomics, version 2.1.0). All subsequent quality control and data processing steps, including removal of doublets, open inferred gene activity, dimensionality reduction, clustering and chromatin peak calling were performed using ArchR (version 1.0.1) ^31^. We used canonical correlation analysis in Seurat (v4.3.0) ^25^ to assign cell type identities to snATAC-Seq clusters through integration with the GE level 1 snRNA-seq data described above, retaining ATAC-Seq data from nuclei that were predicted to derive from progenitor cells or developing neurons of the CGE, LGE or MGE. ATAC-Seq data from retained nuclei were then re-clustered using ArchR and high confidence (FDR < 0.05) open chromatin regions (OCRs) determined in each of the four major cell types using MACS2 ^32^. For a detailed description of snATAC-Seq data processing, see Supplementary Methods.

### Testing enrichment of schizophrenia SNP heritability in OCRs mapped in individual cell populations of the ganglionic eminences

We used SLDSR ^30^ to test enrichment of schizophrenia SNP heritability in OCRs mapped within progenitor cells and developing neurons of the CGE, LGE and MGE. SNPs associated with human height ^29^ were again used as a comparison non-brain phenotype. The hg38 genomic coordinates of OCRs identified in each cell population were converted to hg19 using LiftOver (https://genome.sph.umich.edu/wiki/LiftOver) and enrichment of SNP heritability within OCRs for each cell type was assessed against that of 53 (v1.2) baseline genomic annotations ^30^ and all OCRs identified in all cell populations of the GE in this study. OCRs overlapping the MHC region (hg19; chr6:28,477,797-33,448,354) were removed prior to these analyses. The resulting Z-scores were used to calculate one-tailed enrichment *P*-values for each cell population.

### Mapping fine-mapped schizophrenia-associated SNPs to cell-specific OCRs within the ganglionic eminences

Sets of credible causal variants derived by fine-mapping schizophrenia loci (95% credible sets), were obtained from Supplementary Table 11 of Trubetskoy et al ^16^. These SNPs were converted into hg38 coordinates and mapped to OCRs identified in each broad GE cell population. For fine-mapped SNPs in OCRs located in distal intergenic regions (i.e. not in exons, introns or promoters, the latter defined as within 1000bp upstream and 100bp downstream of the transcription start site), we determined potential gene targets of the SNP by calculating co- accessibility between the intergenic OCR and OCRs located at gene promoters within 100kb of that OCR using ArchR ^31^.

## Results

### Enrichment of schizophrenia common variant risk in genes with high expression specificity for developing neuronal populations of the ganglionic eminences

scRNA-Seq data from the study of Shi and colleagues ^24^ were clustered according to global gene expression profile and cell clusters annotated based on expression of canonical markers and other differentially expressed genes (Figure 1). Clusters annotated as belonging to the same broad level 1 cell-type were aggregated to form 3 clusters of developing post-mitotic neurons, deriving from either the CGE (CGE-N; markers: *CALB2, SCGN, PCDH9, ANKS1B*), LGE (LGE-N; markers: *FOXP1, ZNF503, SERTAD4, ISL1*), or MGE (MGE-N; markers: *LHX6, NXPH1*), and 3 additional clusters corresponding to IPCs (markers: *DLL1*, *DLL3* and *CCND2*), progenitor cells (markers: *HES1, OLIG2, PAX6, GSX2, PTPRZ1*) and microglia (markers: *SPI1, CD68*).

**Figure 1:**
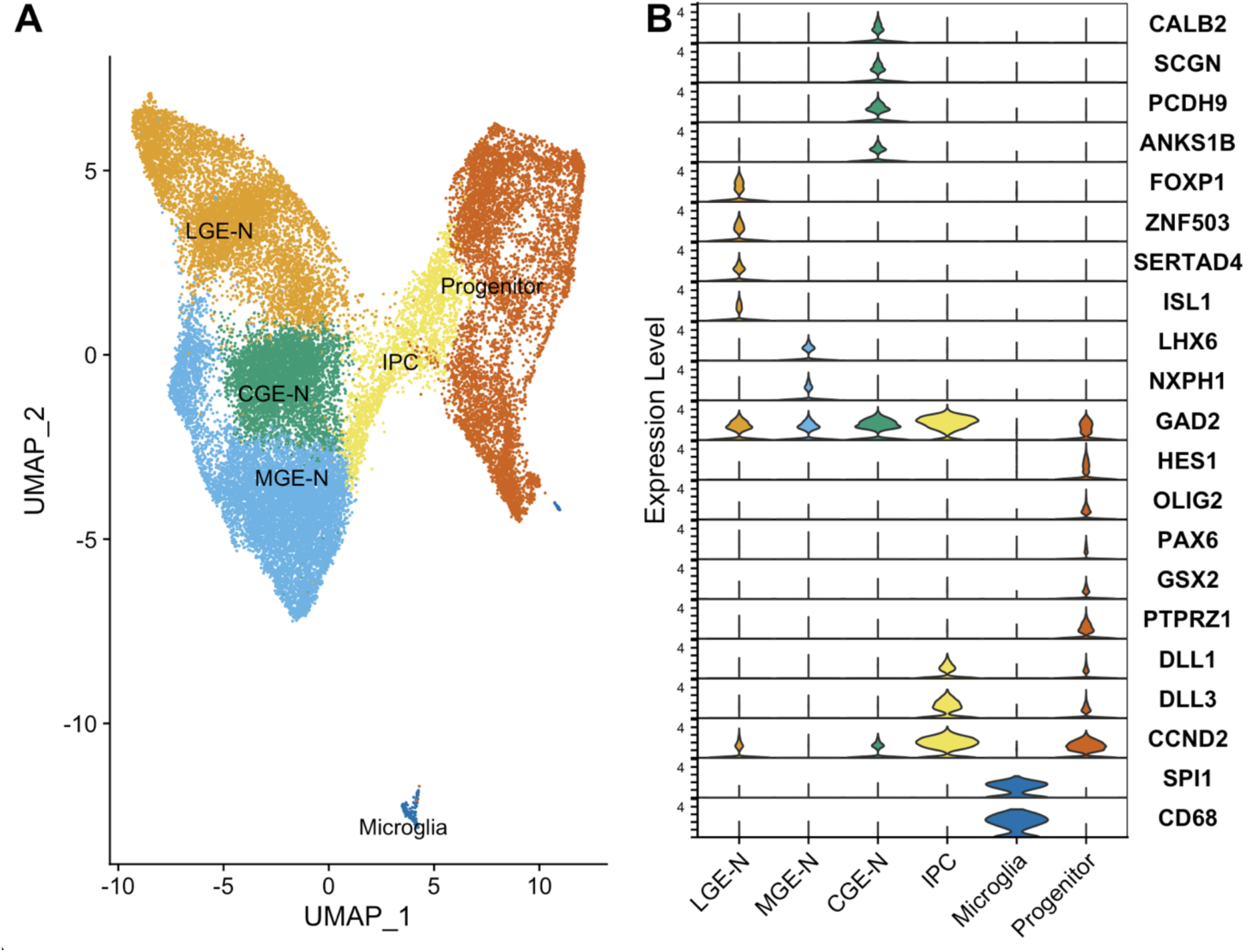
Initial analyses of single-cell RNA sequencing data from the ganglionic eminences. ^24^. Cells were clustered according to gene expression profile using Seurat 4.3.0 ^25^ and visualized in two-dimensional space using UMAP. Cells that were labelled by Shi et al ^24^ as deriving from tissue outside of the GE were excluded prior to clustering. A) Broad ‘Level 1’ cell types: developing neurons from the MGE (MGE-N), developing neurons from the CGE (CGE-N), developing neurons from the LGE (LGE-N), intermediate progenitor cells (IPC), progenitor cells and microglia. B) Violin plots showing cell marker gene expression across level 1 cell types.

We tested enrichment of schizophrenia common variant risk ^16^ in genes in the top decile of expression specificity for each broad ‘level 1’ cell type. Genes with high expression specificity for developing neurons of the CGE, LGE and MGE were enriched for schizophrenia liability at a significance level exceeding the Bonferroni threshold of *P* < 8.3 x 10^-3^ in both the MAGMA and SLDSR tests (Figure 2A). We observed no enrichment of schizophrenia liability in genes with high expression specificity for progenitor cells, IPCs or microglia, consistent with our previous study performed on an independent set of human fetal brain samples ^23^. This pattern of enrichment was maintained when down-sampling the number of cells for each population to match that of the smallest cell population (Figure S1A), or when an equal number of genes were tested for each cell population (Figure S1B). In contrast, height-associated variation was not enriched in genes in the top decile of expression specificity of any post-mitotic neuronal population, with only the SLDSR analysis of genes specific to IPCs reaching the Bonferroni-corrected *P*-value threshold (Figure 2B).

**Figure 2:**
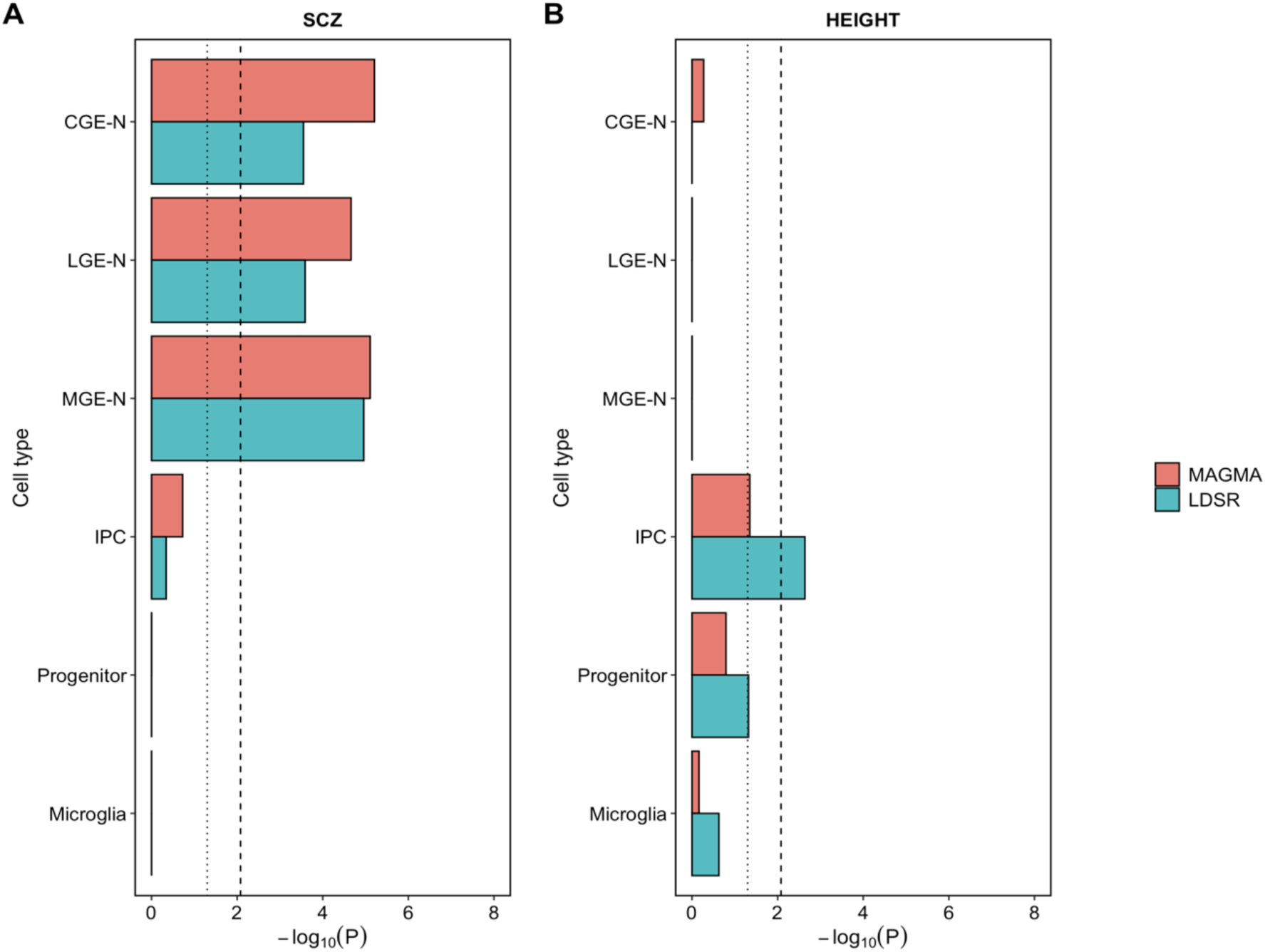
-Log_10_ *P*-values for enrichment of schizophrenia (A) and height (B) common variant associations in genes in the top expression specificity decile of 6 broad level 1 cell types of the ganglionic eminences using MAGMA and SLDSR. The dotted vertical line indicates nominal (*P* < 0.05) significance and the dashed vertical line indicates the Bonferroni-corrected *P*-value threshold for 6 tested cell populations (*P* < 0.0083). CGE-N = developing neurons from the CGE; LGE-N = developing neurons from the LGE; MGE-N = developing neurons from the MGE; IPC = intermediate progenitor cells.

To refine the cell populations of the GE relevant to schizophrenia, we separately sub-clustered developing neurons of the MGE, LGE and CGE (as well as IPCs and progenitor cells) to obtain 36 ‘level 2’ cell populations (Supplementary Figures S2-S6). These comprised 8 developing neuron populations from the LGE, 6 from the MGE, 4 from the CGE, 6 subpopulations of IPCs, 11 subpopulations of progenitor cells and microglia (the latter not sub-clustered due to the low starting number of cells).

As before, we tested genes in the top decile of expression specificity for each of the 36 cell subpopulations for enrichment of schizophrenia common variant liability (Figure 3). Seven level 2 cell types showed significant enrichment exceeding the Bonferroni threshold of *P* < 1.4 X 10^-3^ in both statistical tests. Three of these were predicted to be LGE-derived developing GABAergic medium spiny neuron (MSN) populations destined for the striatum: one predicted to become dopamine D1 receptor-expressing MSNs (LGE-N-1, prominently expressing *ZNF503* and *ISL1*; Supplementary Figure S2B) and two precursors of dopamine D2 receptor-expressing MSNs (LGE-N-2, marker: *CXCL12*; LGE-N-4, marker: *PENK*; Supplementary Figure S2B). Within the MGE, enrichment of schizophrenia common variant associations was observed in genes with high expression specificity for a somatostatin (SST)- positive GABAergic interneuron population (MGE-N-3, markers: *SST*, *MAF*; Supplementary Figure S3B) and a population predicted to develop into cholinergic neurons (MGE-N-2, markers: *LHX8*, *ZIC1*, *CNTNAP2*; Supplementary Figure S3B). Within the CGE, genes with high expression specificity for two calretinin (*CALB2*)- positive sub-populations of developing GABAergic neuron were also enriched for schizophrenia genetic liability (CGE-N-1, marker: *NRIP3*, and CGE-N-2, markers: *BEX2*, *ARL6IP5*, *VSTM2A*; Supplementary Figure S4B). Significant enrichment of schizophrenia genetic liability (exceeding the Bonferroni threshold in both MAGMA and SLDSR) was maintained in all 7 implicated cell populations when an equal number of genes was analysed for each population (Supplementary Figure S7B), and, with the exception of CGE-N-1, when the number of cells from those subpopulations were down-sampled (Supplementary Figure S7A). Schizophrenia associations were not enriched in genes with high expression specificity for any IPC or progenitor cell type. Common variants associated with human height were not enriched in genes with high expression specificity for any cell subpopulation at the Bonferroni-corrected *P*-value threshold.

**Figure 3.**
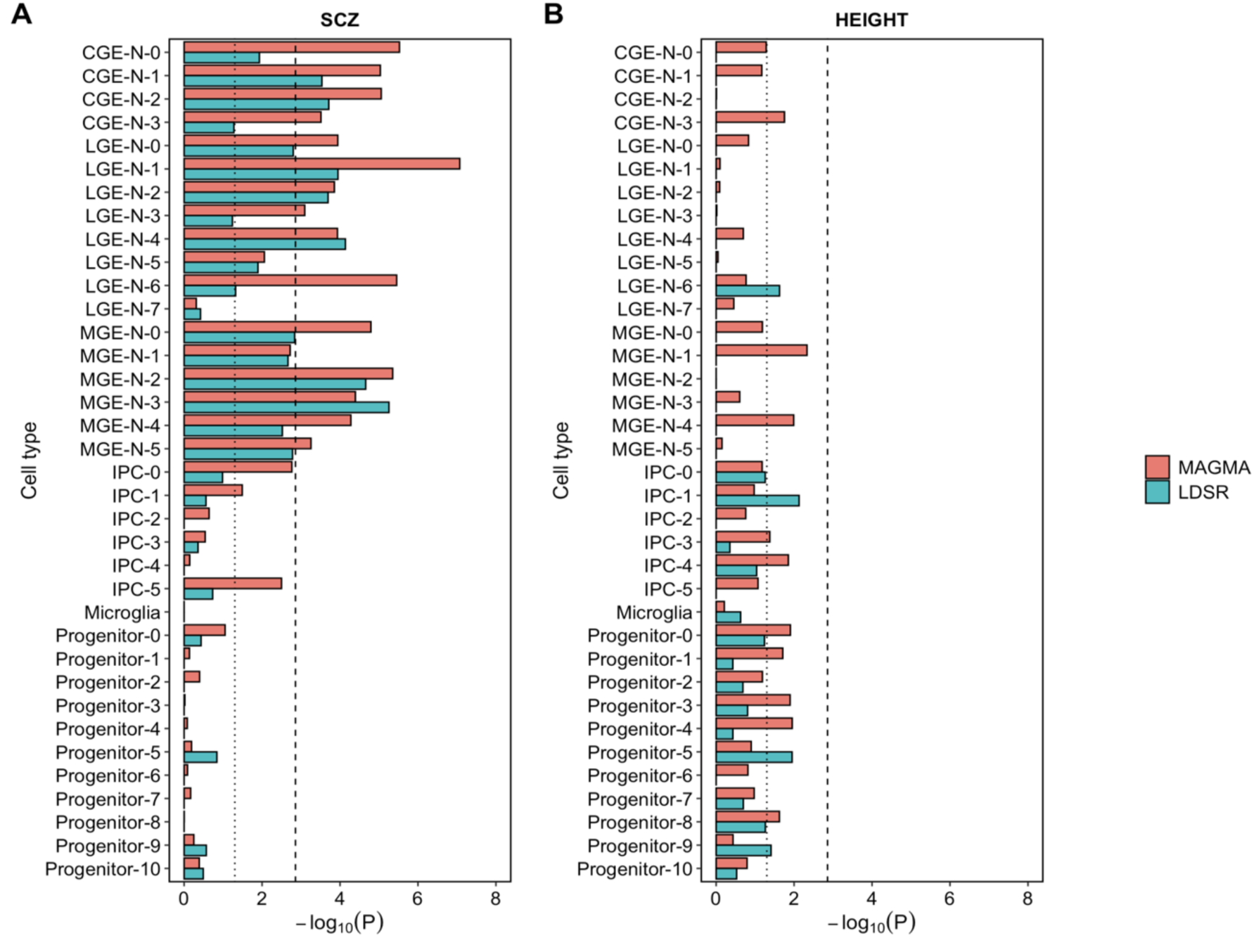
-Log_10_ *P*-values for enrichment of schizophrenia and height common variant associations in genes in the top expression specificity decile of 36 ‘level 2’ ganglionic eminence subpopulations using MAGMA and SLDSR. The dotted vertical line indicates the nominal (*P* < 0.05) significance level and the dashed vertical line indicates the Bonferroni-corrected *P*-value threshold for 36 tested cell populations (*P* < 0.0014). CGE-N = developing neurons from the CGE; LGE-N = developing neurons from the LGE; MGE-N = developing neurons from the MGE; IPC = intermediate progenitor cells.

Given that all cell populations that are enriched for schizophrenia genetic associations are developing neurons, we tested for signal independence between the 7 implicated sub-populations through conditional analyses. After separately conditioning the genes in the top expression specificity decile for each implicated cell type on genes in the top decile of each of the 6 other cell types, all 7 developing neuron populations remained significantly (*P* < 0.05) enriched for genetic association with schizophrenia (Supplementary Figure S8). To determine whether enrichments were also independent of those observed in GABAergic neuron populations of the adult brain implicated in schizophrenia susceptibility through the same methods (mature interneurons of the cerebral cortex and medium spiny neurons of the striatum) ^16^, we similarly performed MAGMA and SLDSR on implicated fetal cell types conditioning on genes in the top specificity decile for each of the two adult neuron populations. Again, all implicated cell populations of the GE remained significantly enriched for association with schizophrenia (all *P* < 0.05; Supplementary Figure S9). Thus, all 7 level 2 neuron populations of the GE show independent evidence for association with schizophrenia, and, moreover, these associations are at least partly independent of those observed in adult GABAergic cells using the same methods and GWAS dataset.

### Single nuclei open chromatin profiling of the human ganglionic eminences

The GWAS variants that are most strongly associated with schizophrenia predominantly reside in non-coding regions of the genome ^16^ and are therefore likely to increase risk for the disorder through effects on gene regulation. Active regulatory genomic regions are associated with accessible, or ‘open’, chromatin, which exposes the DNA to transcription factors and other modulators of gene expression. To map open chromatin regions (OCRs) within cell populations of the GE (and thus determine the schizophrenia-associated SNPs that may be operating within them), we performed snATAC-Seq on whole GE dissected from 3 karyotypically normal fetuses from the second trimester of gestation. After strict quality control, we retained snATAC-Seq data from 7,306 nuclei (Supplementary Figure S10); these were further refined to 3,163 nuclei that we were able to confidently label as deriving from developing neuron or progenitor cells of the GE based on integration with the level 1 scRNA-Seq data of Shi and colleagues ^24^ (Figure 4A). Following re-clustering, we confirmed the identity of aggregated nuclei based on elevated gene scores for regional markers (Figure 4B).

**Figure 4.**
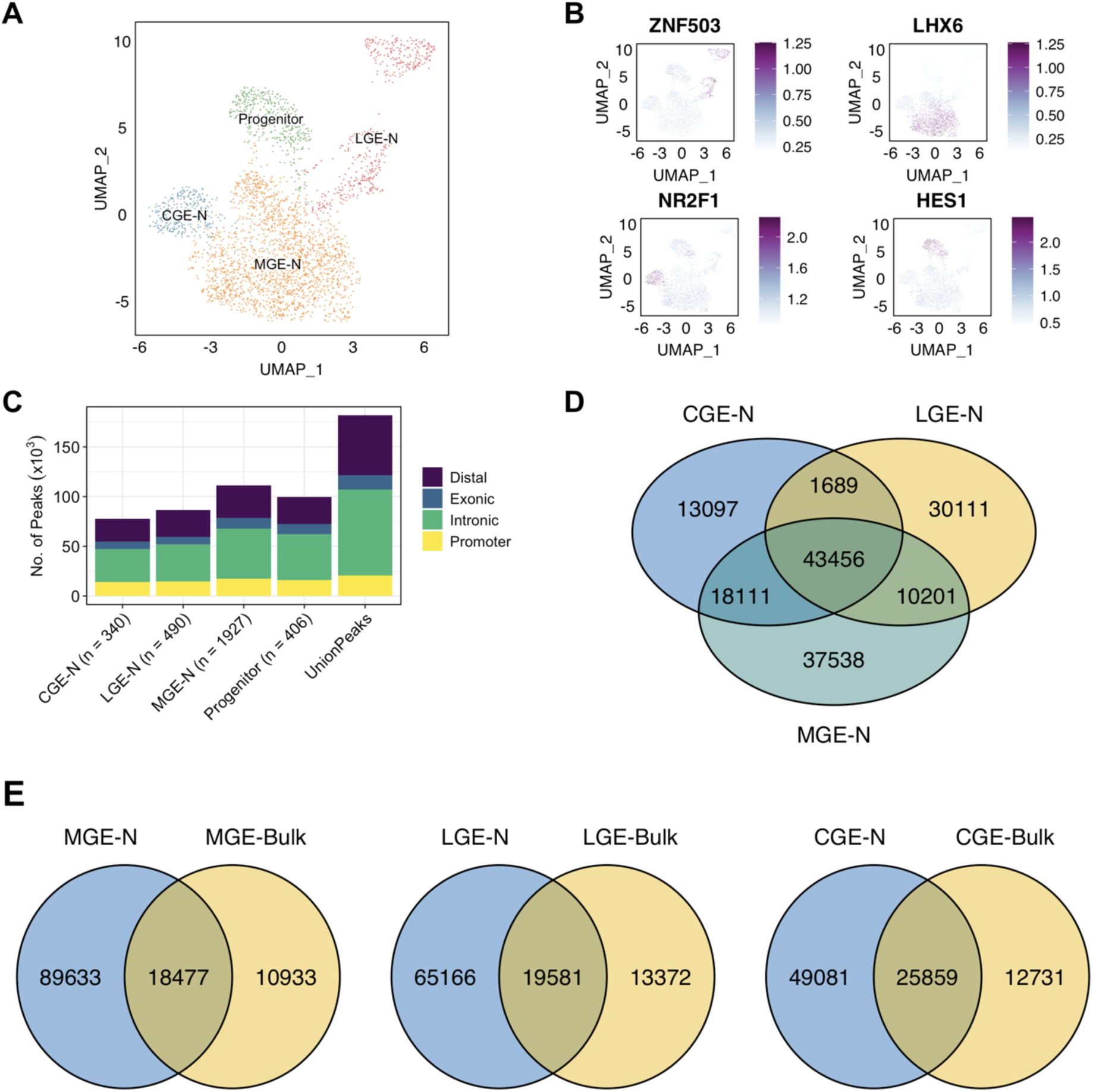
Single nuclei ATAC-Seq of the human ganglionic eminences. A) snATAC-Seq clusters after integration with scRNA-Seq data from the study of Shi et al ^24^. Nuclei were clustered according to open chromatin profile using ArchR ^31^ and visualized in 2D space using Uniform Manifold Approximation and Projection (UMAP) ^25^. CGE-N = developing neurons from the CGE; LGE-N = developing neurons from the LGE; MGE-N = developing neurons from the MGE. B) Clustered nuclei were classed as deriving from major cell types based on gene scores (inferred gene expression based on chromatin accessibility at the gene locus) for known marker genes. C) Number of open chromatin regions identified in each cell population according to genomic annotation (promoter sequence is arbitrarily defined as within 1000 bp upstream and 100 bp downstream of a transcription start site). D) Overlap of open chromatin regions identified in developing neurons of the CGE, LGE and MGE. E) Overlap between open chromatin regions identified in developing neurons of the MGE, LGE and CGE and those identified through ATAC-Seq of bulk tissue from those regions by Markenscoff-Papadimitriou et al ^33^.

We identified between 77,465 and 111,210 high confidence (FDR < 0.05) OCRs within each level 1 cell population of the GE (Figure 4C; Supplementary Tables S3 – S6). Identified OCRs were concentrated in known promoters (defined as between 1000 bp upstream and 100 bp downstream of a TSS) and introns, with approximately a third in distal intergenic regions (Figure 4C). OCRs were highly enriched for transcription factor binding motifs appropriate for their region and cell type (Supplementary Figure S11). Although developing neurons of the CGE, LGE and MGE share a large proportion of their OCRs, we identified between 13,097 and 37,538 OCRs unique to each region (Figure 4D). Less than 35% of the OCRs identified in developing neurons of the MGE, CGE and LGE had been previously identified using ATAC-Seq in bulk tissue dissected from these regions of the human fetal brain ^33^ (Figure 4E), highlighting the value of single nuclei ATAC-Seq in capturing additional, cell-specific OCRs.

### Enrichment of schizophrenia SNP heritability in OCRs identified in developing neurons of the ganglionic eminences

To complement our snRNA-Seq based enrichment analyses, we used SLDSR to test for enrichment of schizophrenia SNP heritability in the OCRs mapping to each level 1 GE cell population. Consistent with our analyses based on cell-specific gene expression (and a role for altered fetal gene regulation in mediating genetic risk for schizophrenia ^34–36^), SNP heritability for schizophrenia was highly enriched in OCRs mapped within developing neuron populations of the CGE, LGE and MGE but not in OCRs mapped in progenitor cells (Figure 5A). SNP heritability for human height was not enriched in OCRs mapped to any developing neuron population of the GE, but was significantly enriched in OCRs mapped to progenitor cells (Figure 5B).

**Figure 5:**
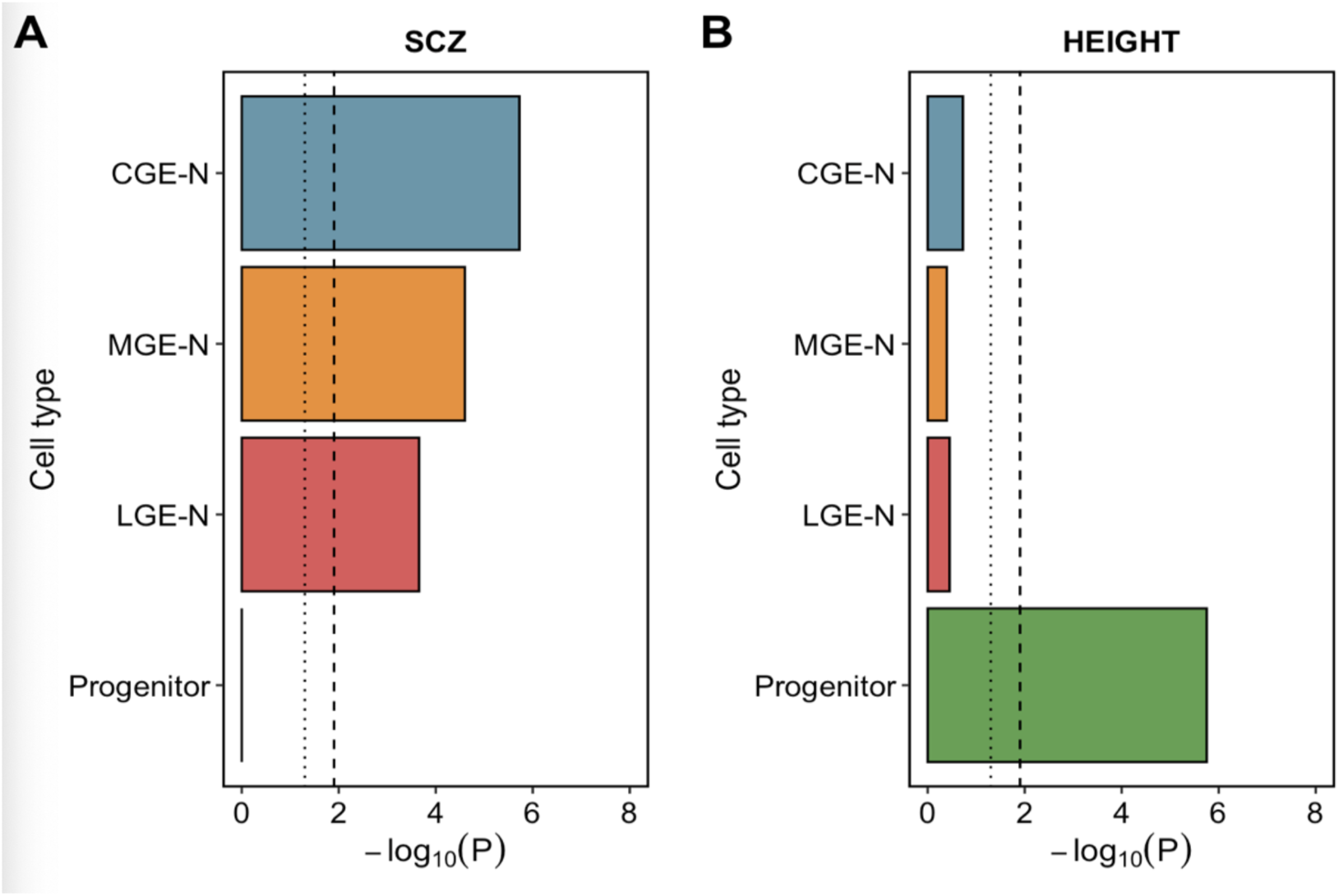
-Log_10_ *P*-values for enrichment of SNP heritability for schizophrenia and height in OCRs mapped within individual cell populations of the ganglionic eminences. Analyses were performed using SLDSR ^30^; enrichment *P-*values were derived from Z-scores accounting for 53 (v1.2) baseline genomic annotations ^30^ and OCRs identified in all other cell populations of the GE. The dotted vertical line indicates nominal (*P* < 0.05) significance and the dashed vertical line indicates the Bonferroni-corrected *P*-value threshold for 4 tested cell populations (*P* < 0.0125). CGE-N = developing neurons of the CGE; LGE-N = developing neurons of the LGE; MGE-N = developing neurons of the MGE.

Ziffra and colleagues ^37^ used snATAC-Seq to map OCRs within cell populations of the fetal cerebral cortex, reporting enrichment of schizophrenia SNP heritability in OCRs overlapping enhancer marks in excitatory neuron populations (as well as cortical interneurons predicted to derive from the CGE). We assessed the extent to which enrichments of schizophrenia SNP heritability in OCRs mapped within developing neuron populations of the GE are independent of those in OCRs mapped in developing glutamatergic neurons by including the latter in the SLDSR analyses (Supplementary Figure S12). Schizophrenia SNP heritability remained significantly enriched (above the Bonferroni-corrected *P*-value threshold) in OCRs mapped in each of the neuronal GE populations when OCRs mapped in developing glutamatergic neurons of the fetal cerebral cortex were accounted for. These findings provide further evidence for a distinct contribution of early GE-derived neuronal development in susceptibility to schizophrenia.

### Localization of fine-mapped schizophrenia-associated SNPs in OCRs mapped within cell populations of the ganglionic eminences

A limitation of GWAS is that the causal genetic variants underlying associations are usually unclear. This is in part due to linkage disequilibrium (which often results in multiple variants at a locus displaying similar levels of association) but also reflects limited functional characterization of the noncoding genome. Trubetskoy and colleagues ^16^ used Bayesian fine-mapping ^38^ to determine the posterior probabilities of individual SNPs at genome-wide significant risk loci for schizophrenia being causal and to determine credible sets of potentially causal SNPs at each locus. We further refined the 20,766 SNPs that collectively capture 95% of the posterior probability of being causal at each of the 255 fine-mapped genome-wide significant loci for schizophrenia ^16^ according to whether they are located within OCRs mapped within cell populations of the GE. We list all fine-mapped SNPs located within GE cell OCRs in Supplementary Table S7. These include non-coding SNPs located within 24 genes that were prioritized as candidates by Trubetskoy et al ^16^, such as the intronic SNP rs2944819 in *CALN1* (located within an OCR mapped in CGE-N and LGE-N) and the intronic SNP rs11972718 in *NXPH1* (located within an OCR mapped only in MGE-N). For fine-mapped schizophrenia-associated SNPs in distal intergenic OCRs, we sought to identify genes that they putatively regulate through co-accessibility with promoter OCRs using ArchR ^31^. Accessibility at 7 distal intergenic OCRs containing fine-mapped SNPs correlated significantly with promoter OCR accessibility for genes in the region across cell types (Supplementary Table S8). These include a distal OCR (containing SNP rs62183854) that is co-accessible with promoter OCRs for *DLX1* (correlation = 0.64; FDR = 1.28 X 10^-47^) and *DLX2* (correlation = 0.56; FDR = 3.76 X 10^-34^), transcription factors with important roles in GABAergic neuron development ^39^.

## Discussion

A rational approach for identifying cell populations that are etiologically relevant to a trait of interest is to integrate genomic data for that trait with cell-specific functional annotations ^20–22,40^. In this study, we combined data from the largest GWAS of schizophrenia ^16^ with high resolution scRNA-Seq data from the human GE ^24^ to implicate defined populations of developing GABAergic and cholinergic neuron in common variant liability to schizophrenia. We complement this work by performing snATAC-Seq on fetal brain tissue to map potential regulatory genomic regions in individual cell populations of the human GE. Consistent with our scRNA-Seq-based analyses, we find that SNP heritability for schizophrenia is strongly enriched in predicted regulatory genomic regions operating in developing GE neurons, and we further use these data to functionally prioritize non-coding SNPs associated with the disorder.

As well as supporting the hypothesis that schizophrenia has an early neurodevelopmental component ^41,42^, our findings provide evidence for a primary GABAergic disturbance in the disorder ^9–11,43,44^. One of the developing GABAergic cell types we implicate in schizophrenia genetic liability is SST-positive interneurons of the MGE. Multiple post-mortem studies have reported decreased expression of SST in the cerebral cortex of people with schizophrenia ^7,8,45^, with SST reductions also observed in hippocampus ^46^ and amygdala ^47^. Our findings suggest that this could partly reflect a developmental vulnerability in these neurons. Enrichment of schizophrenia genetic liability was also observed in genes with high expression specificity for calretinin (*CALB2*)- positive subpopulations of developing GABAergic neuron from the CGE. Although calretinin expression is reported to be unchanged in the prefrontal cortex in schizophrenia ^6,48–50^, it has been found to be reduced in the caudate nucleus in the disorder ^51^. We also found enrichment of schizophrenia common variant liability in genes with high expression specificity for 3 populations of developing GABAergic MSN populations destined for the striatum. MSNs constitute the majority of striatal neurons ^52,53^ and can be broadly subdivided into those expressing the dopamine D1 or D2 receptors. We implicate in schizophrenia susceptibility one subpopulation that is predicted (based on known cell-type markers ^24^) to become D1 receptor-expressing MSNs and two subpopulations that are predicted (based on known cell-type markers ^24^) to develop into D2 receptor-expressing MSNs. Schizophrenia common variant liability has also been found to be enriched within genes with high expression specificity for striatal MSNs from the adult mouse ^16,20,22^ and in genes marking several D1 receptor- and D2 receptor- expressing MSN populations of the adult human ventral striatum (nucleus accumbens) ^53^. Given that the dopamine D2 receptor is a primary target of antipsychotic medication ^54^, D2 receptor-expressing MSNs are plausibly initial mediators of the therapeutic response to these drugs ^55^. Our data, along with those from MSNs of the adult striatum ^16,20,22,53^, suggest that this may compensate for an abnormality in these neurons in schizophrenia. In addition, we provide evidence that abnormalities extend to certain D1-receptor expressing MSNs, consistent with recent data suggesting these to be important mediators of antipsychotic drug efficacy ^56^.

Our finding that a population of developing LHX8-positive neurons of the MGE is also enriched for genetic association with schizophrenia suggests a role for cholinergic neuron development in later susceptibility to the condition. The MGE gives rise to cholinergic interneurons of the striatum as well as basal forebrain cholinergic projection neurons, the former modulating GABAergic neuron activity within the striatum, while the latter project to the cerebral cortex, hippocampus and amygdala ^4^. Dysfunction of either of these cell types could plausibly contribute to schizophrenia symptomatology ^57,58^; indeed, activators of muscarinic acetylcholine receptors are currently in clinical development as novel treatments for the disorder ^59^.

As well as providing independent evidence for a role of GE-derived neuron development in genetic predisposition to schizophrenia, our snATAC-Seq data enabled us to define potential regulatory regions of the genome operating within individual cell populations of the GE with which to functionally prioritise schizophrenia-associated SNPs. Such information could be used in the design of functional assays to determine direct effects of these variants on gene expression ^60^ and cellular function ^61^. In addition, regulatory genomic annotations have been shown to significantly improve the *trans*-ancestry portability of polygenic risk scores ^62^ and will be important in interpreting the findings of whole genome sequencing studies of human disorders. We provide the genomic coordinates of identified cell-specific OCRs of the GE (Supplementary Tables S3-S6) towards such efforts.

Our finding that schizophrenia common variant genetic liability is enriched in neurons rather than progenitor cells is consistent with other studies using single cell / nuclei sequencing data from the developing human brain ^23,37^ and the view that schizophrenia is primarily a neuronal disorder ^63^. We ^23^ and others ^37^ have found particularly strong enrichment of schizophrenia common variant genetic risk in developing glutamatergic neurons of the prenatal brain, although those and the present findings indicate an independent contribution of GABAergic neuron development. A limitation of our study is that we were not able to test enrichment of schizophrenia genetic risk in parvalbumin-positive GABAergic interneurons, which have been strongly implicated in schizophrenia through post-mortem studies ^6–8^, as parvalbumin is not expressed in these neurons until after birth ^64–66^. Further insight into the cellular etiology of schizophrenia will be provided by single cell datasets from the human brain that incorporate a wider range of developmental stages and brain regions, as well as spatial information ^67^.

## Acknowledgements

The human fetal material was provided by the Joint MRC / Wellcome (MR/R006237/1, MR/X008304/1 and 226202/Z/22/Z) Human Developmental Biology Resource (http://www.hdbr.org). We thank Dr. Joanne Morgan for assistance with sequencing.

## Data availability

Gene expression cell specificity values and the genomic coordinates of all open chromatin regions identified in this study are provided in Supplementary Tables.

