## Supplementary Methods for "Genetic implication of prenatal GABAergic and cholinergic neuron development in susceptibility to schizophrenia"

snATAC-Seq data preparation

snATAC-Seq quality control and data processing steps were performed using the chromatin accessibility software suite ArchR, version 1.0.1[1]. Default settings were used for all ArchR functions unless otherwise stated. A tile by cell matrix containing sequencing fragment counts tallied in 500bp-wide non-overlapping genome-wide bins (or tiles) was created for each sample (ArchR::createArrowFiles). The Y chromosome, mitochondrial genome and ENCODE Data Analysis Center exclusion list regions[2] were excluded from this matrix. A gene score matrix was also generated containing inferred gene expression scores for each cell based on chromatin accessibility within the gene body and distal regions within 100kb up- and down-stream of the gene start-site (unless overlapping another gene), weighted by genomic distance. Cells were excluded from the analysis if fewer than 3000 unique high-quality nuclear reads were captured and/or the transcription start site (TSS) enrichment score for that cell was not greater than 4. Doublets were then identified and removed on a per sample basis (ArchR:: addDoubletScores). To reduce the dimensionality of the data *ArchR::addIterativeLSI* was run. Nuclei for all samples were then clustered, and clusters were visualised in 2D space via UMAP (*ArchR::addClusters* and *ArchR::addUMAP* respectively).

As snATAC-Seq data are sparse (mostly 0s) and binary (a genomic locus is either accessible or not in a single cell), it is not possible to perform traditional bulk ATAC-Seq analyses, such as peak calling, on data from individual nuclei with statistical confidence. As such, data aggregation methods are employed in snATAC-Seq studies to facilitate downstream analyses. In order to call peaks on the snATAC-Seq data, we ran the *ArchR::addGroupCoverage* function, which generates a set of pseudo-bulk replicates for each cluster using a tiered priority method. A pseudo-bulk replicate contains aggregated fragment data that has been taken from a sampled group of cells from a single cluster. Peaks were then called on the cluster-specific pseudo-bulk replicates using *ArchR::addReproduciblePeakSet*. This function uses an iterative overlap peak merging process where peaks are first called on each pseudo-bulk replicate individually, using MACS2[3], and then they are transformed into peaks of fixed width (501bps). The final output of this process was a single merged peak set for each cell type consisting of non-overlapping 501bp-wide genomic loci (peaks).

To assign cell type identities to the snATAC-Seq cell populations, canonical correlation analysis (CCA) was used to integrate the snRNA-Seq (see Methods section) and snATAC-Seq datasets. For this, the snATAC-Seq ArchR object was first converted into a Seurat object using the package ArchRtoSignac[4] then CCA was run using Seurat[5] (*Seurat::FindTransferAnchors*). CCA projects the snRNA-seq gene expression values and the snATAC-seq gene score values onto a shared feature space and identifies corresponding pairs of cells, termed 'anchors', between the datasets. These anchors were then leveraged to a create a set of prediction scores for each cell in the query (snATAC-Seq) dataset that measured how similar each query cell was to each class of cell in the reference (snRNA-Seq) dataset. The reference cell class with the maximum prediction score was then mapped onto each query cell (*Seurat::TransferData*). Once the maximum prediction scores and the predicted cell identities for each snATAC-Seq cell was established, these annotations were transferred back onto the original snATAC-Seq ArchR object and only cells with a maximum prediction score of >= 0.5 were retained. Any cells that mapped to excitatory neurons were removed as these were likely cells captured from surrounding cortex during tissue dissection. A small population of oligodendrocyte precursor cells was also removed. Finally, the steps from dimensionality reduction through to peak calling, described above, were run again.

Intersection between ganglionic eminence OCRs

To determine the number of overlapping OCRs shared between the ganglionic eminence neuronal populations (CGE-N, LGE-N, MGE-N) the *ChiPpeakAnno::findOverlapsOfPeaks* function[6] (version 3.26.4) was used, with the minimum peak overlap set to 100bp. OCRs derived from bulk tissue that had been dissected from fetal CGE, LGE and MGE were obtained from supplementary table 2B of the Markenscoff-Papadimitriou et al study[7] . Overlaps between bulk tissue OCRs and regionally equivalent single-nuclei derived OCRs generated in this study (specifically the hg19 aligned OCRs that were lifted over from hg38 during the snATAC-Seq SLDSR analyses; see manuscript), were acquired by running the findOverlapsOfPeaks function again with the same parameters as outlined above. Venn diagrams were generated using the VennDiagram package[8].

Transcription factor motif enrichment analysis

Transcription factor binding motif enrichment in ganglionic eminence OCRs was assessed using the R package monaLisa[9]. First position weight matrices for human transcription factor binding sites were downloaded from the JASPAR2020 database[10] by running TFBSTools::getMatrixByID[11] and hg38 sequence information for all OCRs were obtained using the function Biostrings::getSeq (https://bioconductor.org/packages/Biostrings). Motif enrichment was then tested separately in OCRs from each of the 4 major GE cell types by running monaLisa::calcBinnedMotifEnrR, and setting the background parameter to 'genome' to use a randomly sampled, similarly sized set of sequences as the background set of intervals, accounting for differences in GC content and k-mer composition.
