## Supplementary Figures for "Genetic implication of prenatal GABAergic and cholinergic neuron development in susceptibility to schizophrenia"

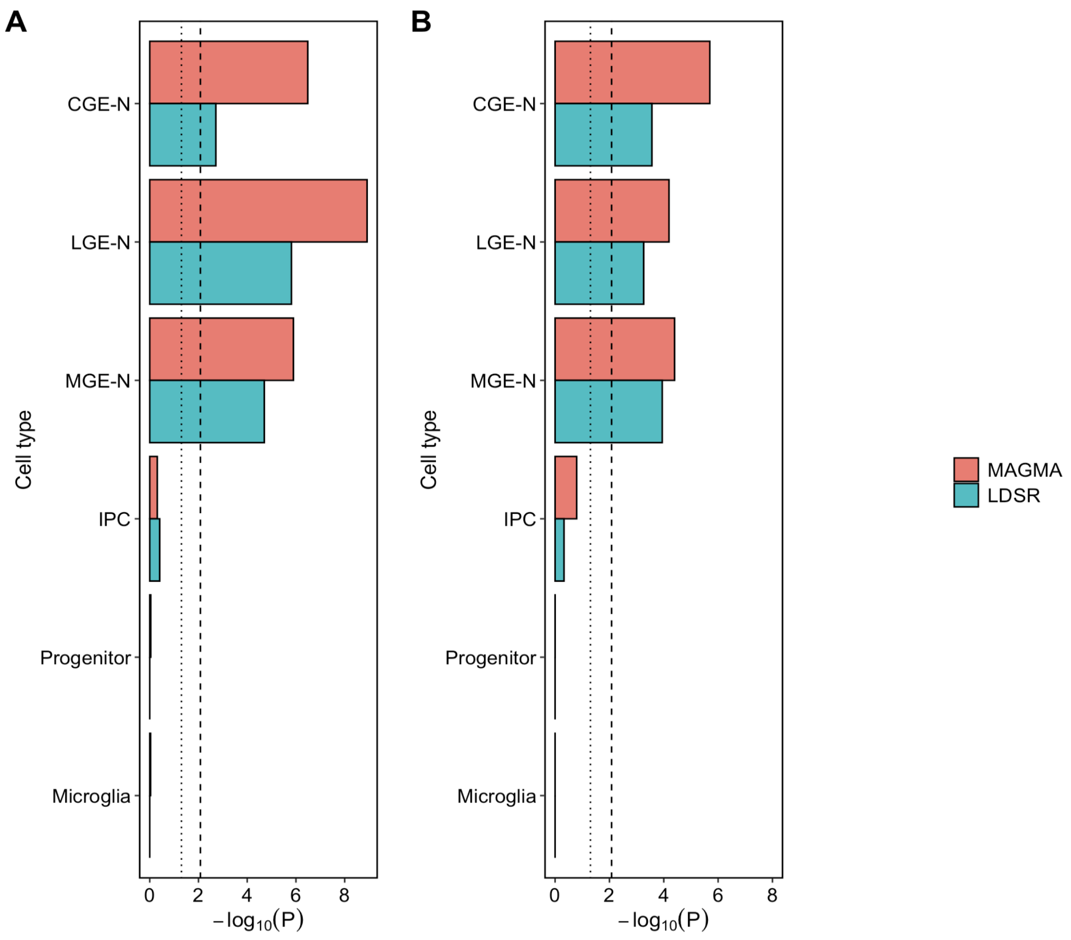


**Figure S1: -Log10 *P*-values for enrichment of schizophrenia genetic associations in genes with high expression specificity for level 1 cell types of the ganglionic eminences using MAGMA and SLDSR after (A) downsampling the number of cells and (B) standardizing the number of genes included for each cell population.** A) -Log10 *P*-values when all level 1 cell types have been down-sampled to match the cell population with the lowest number of cells (microglia, with 242 cells). B) -Log10 *P*-values when the 1000 genes with the highest specificity scores for each cell population are analysed. The dotted vertical line indicates nominal ($P < 0.05$) significance and the dashed vertical line indicates the Bonferroni-corrected *P*-value threshold for 6 tested cell populations at level 1 ($P < 0.0083$).

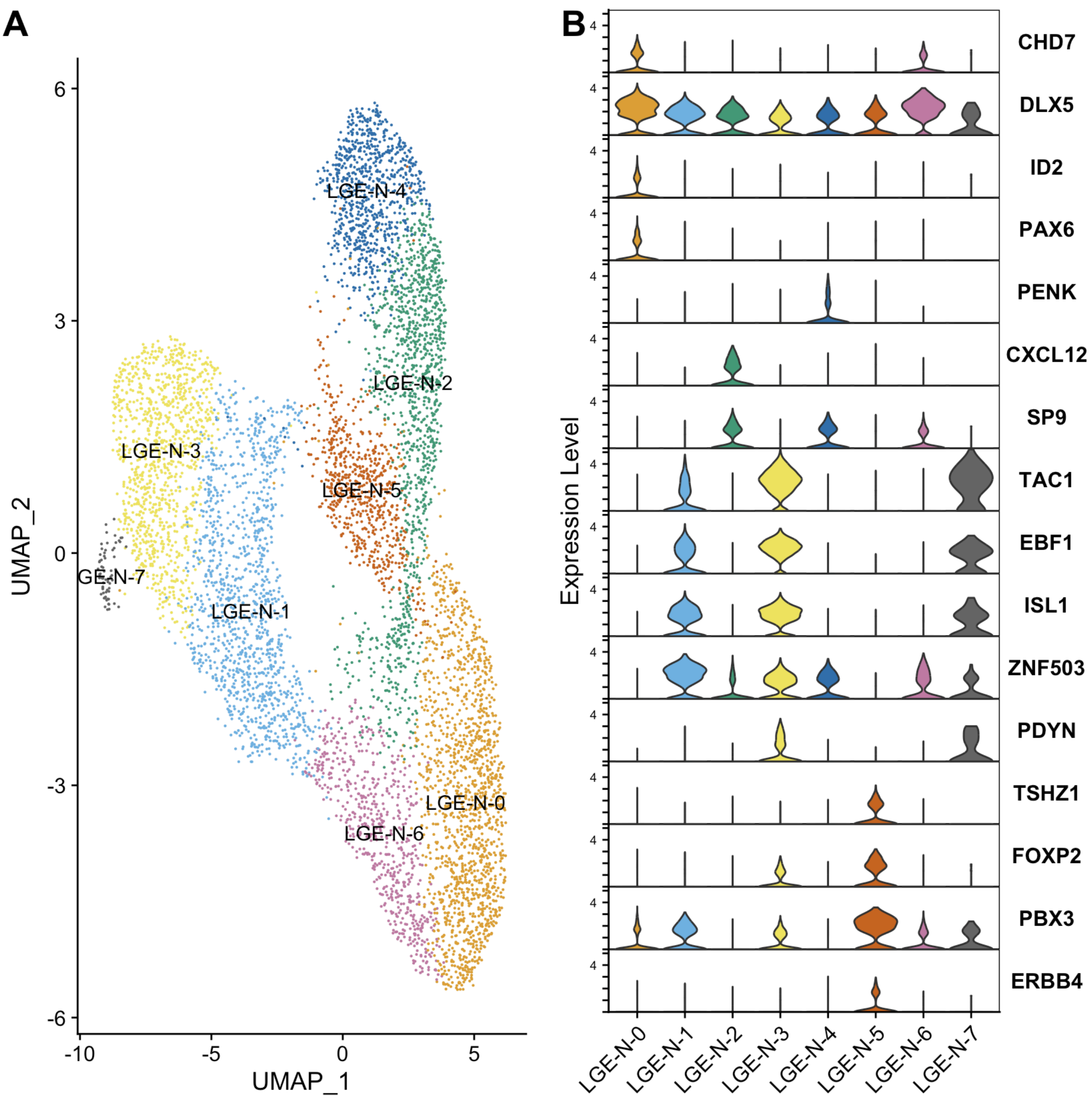


**Figure S2: Subclustering of single-cell RNA-Seq data from developing neurons of the lateral ganglionic eminence.** Cells were clustered according to gene expression profile using Seurat 4.3.0 and visualized in two-dimensional space using UMAP. A) Clusters for level 2 lateral ganglionic eminence neuron (LGE-N) populations. B) Violin plots showing cell marker gene expression across the 8 level 2 lateral ganglionic eminence neuron (LGE-N) subpopulations.

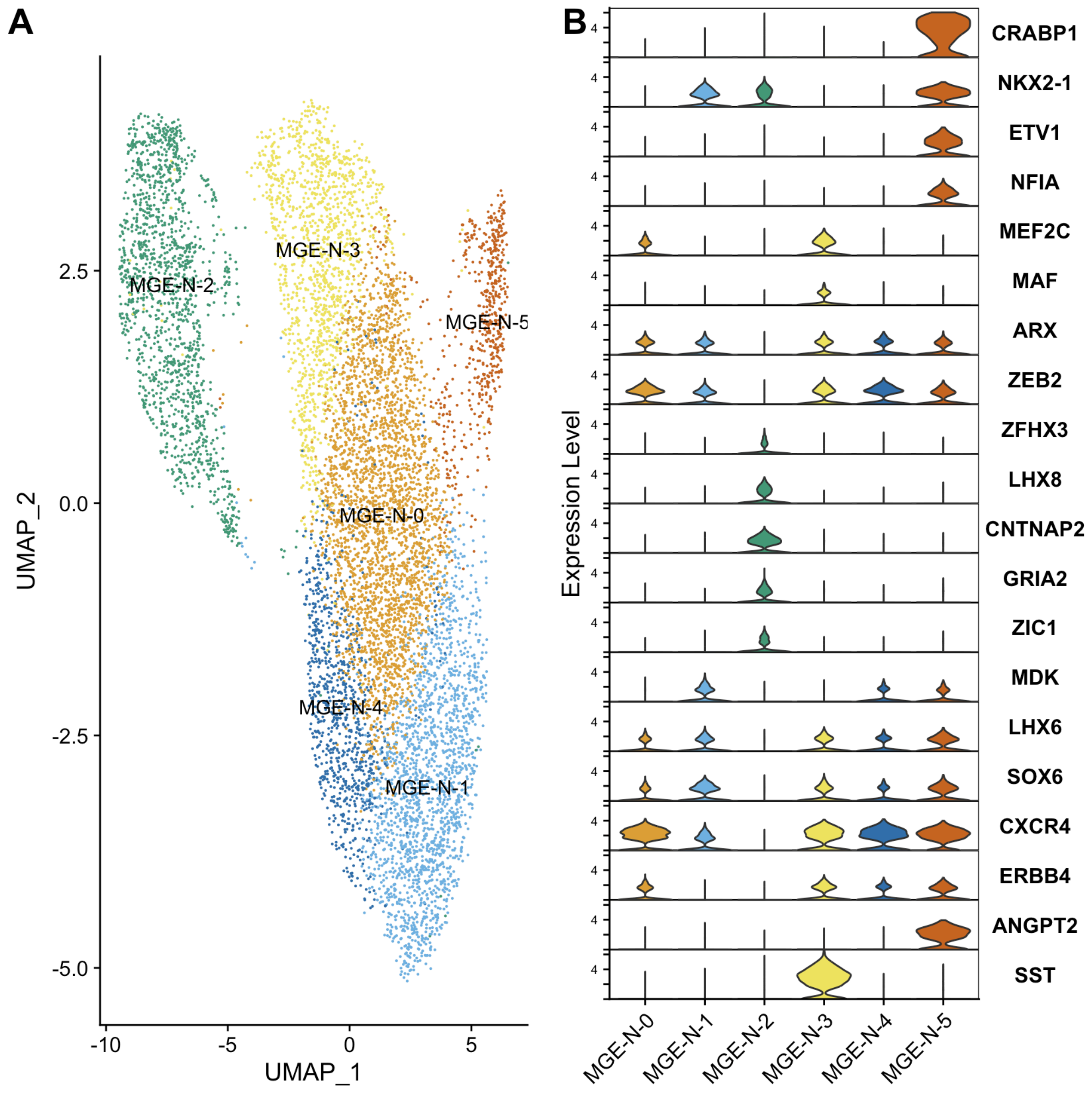


**Figure S3: Subclustering of single-cell RNA-Seq data from developing neurons of the medial ganglionic eminence.** Cells were clustered according to gene expression profile using Seurat 4.3.0 (Hao et al, 2021) and visualized in two-dimensional space using UMAP. A) Clusters for level 2 medial ganglionic eminence neuron (LGE-N) populations. B) Violin plots showing cell marker gene expression across the 6 level 2 medial ganglionic eminence neuron (MGE-N) subpopulations.

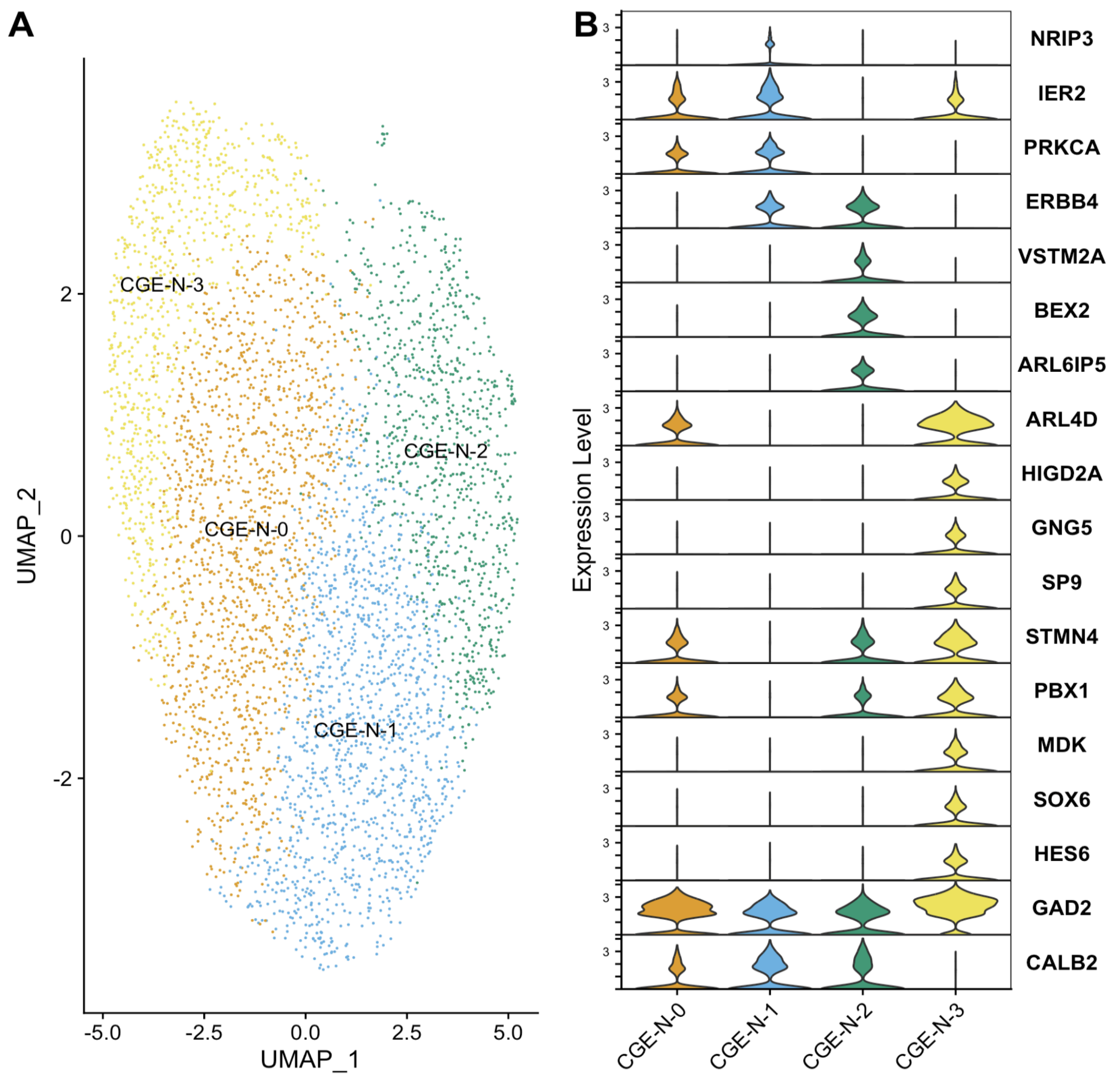


**Figure S4: Subclustering of single-cell RNA-Seq data from developing neurons of the caudal ganglionic eminence.** Cells were clustered according to gene expression profile using Seurat 4.3.0 (Hao et al, 2021) and visualized in two-dimensional space using UMAP. A) Clusters for level 2 caudal ganglionic eminence neuron (CGE-N) populations. B) Violin plots showing cell marker gene expression across the 4 level 2 caudal ganglionic eminence neuron (CGE-N) subpopulations.

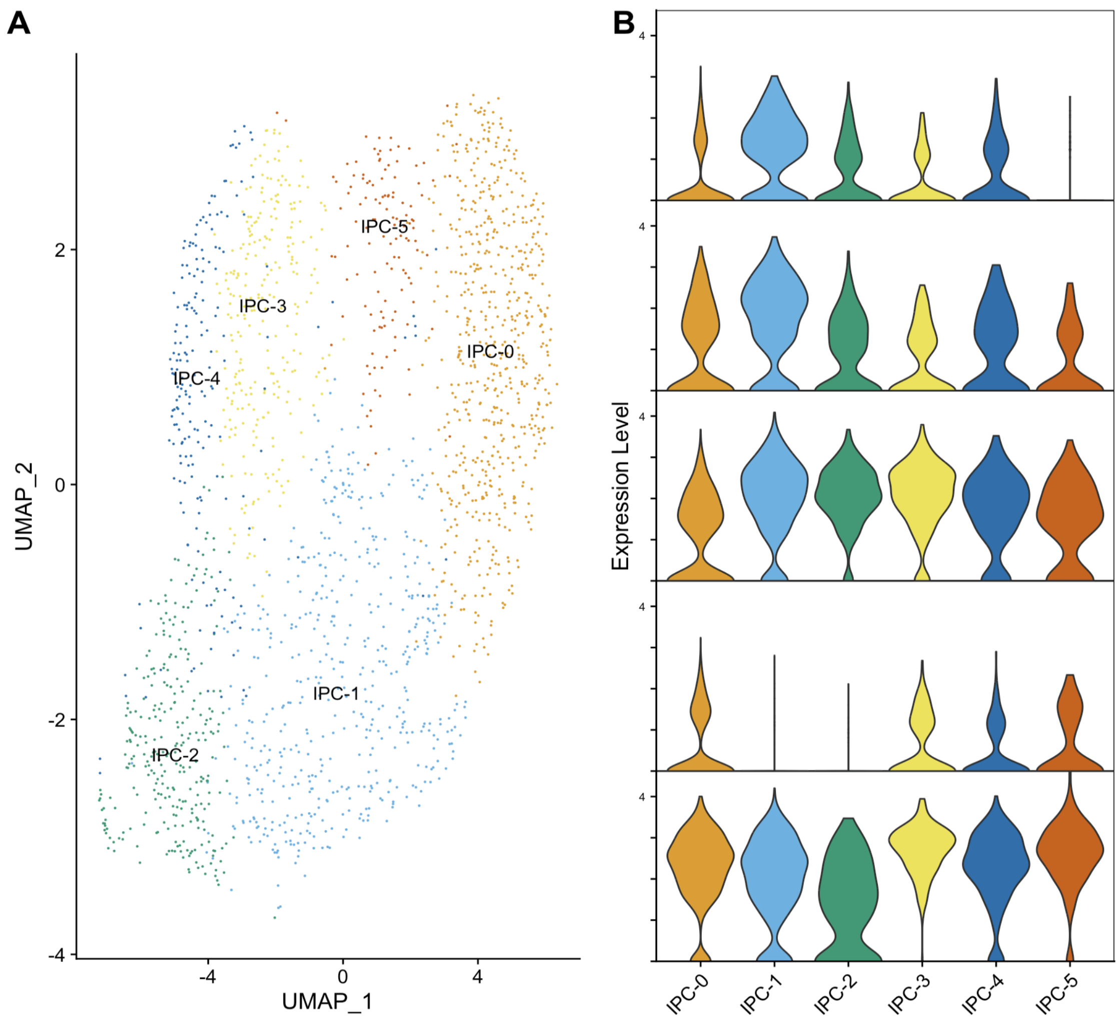


**Figure S5: Subclustering of single-cell RNA-Seq data from intermediate progenitor cells (IPC) of the ganglionic eminences.** Cells were clustered according to gene expression profile using Seurat 4.3.0 (Hao et al, 2021) and visualized in two-dimensional space using UMAP. A) Clusters for level 2 ganglionic eminence IPC populations. B) Violin plots showing cell marker gene expression across the 6 level 2 ganglionic eminence IPC subpopulations.

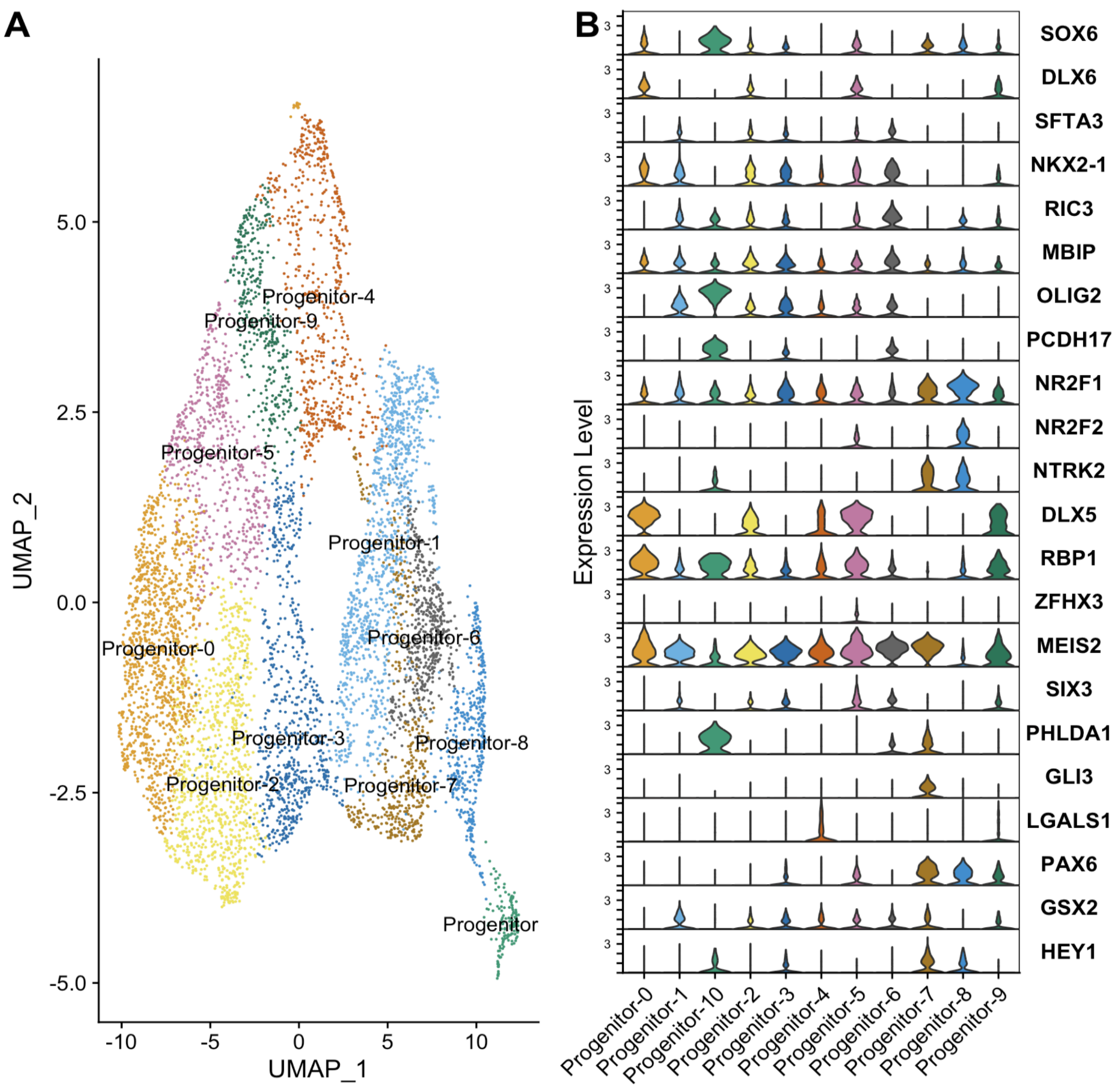


**Figure S6: Subclustering of single-cell RNA-Seq data from progenitor cells of the ganglionic eminences.** Cells were clustered according to gene expression profile using Seurat 4.3.0 (Hao et al, 2021) and visualized in two-dimensional space using UMAP. A) Clusters for level 2 ganglionic eminence progenitor cell populations. B) Violin plots showing cell marker gene expression across the 11 level 2 ganglionic eminence progenitor cell subpopulations.

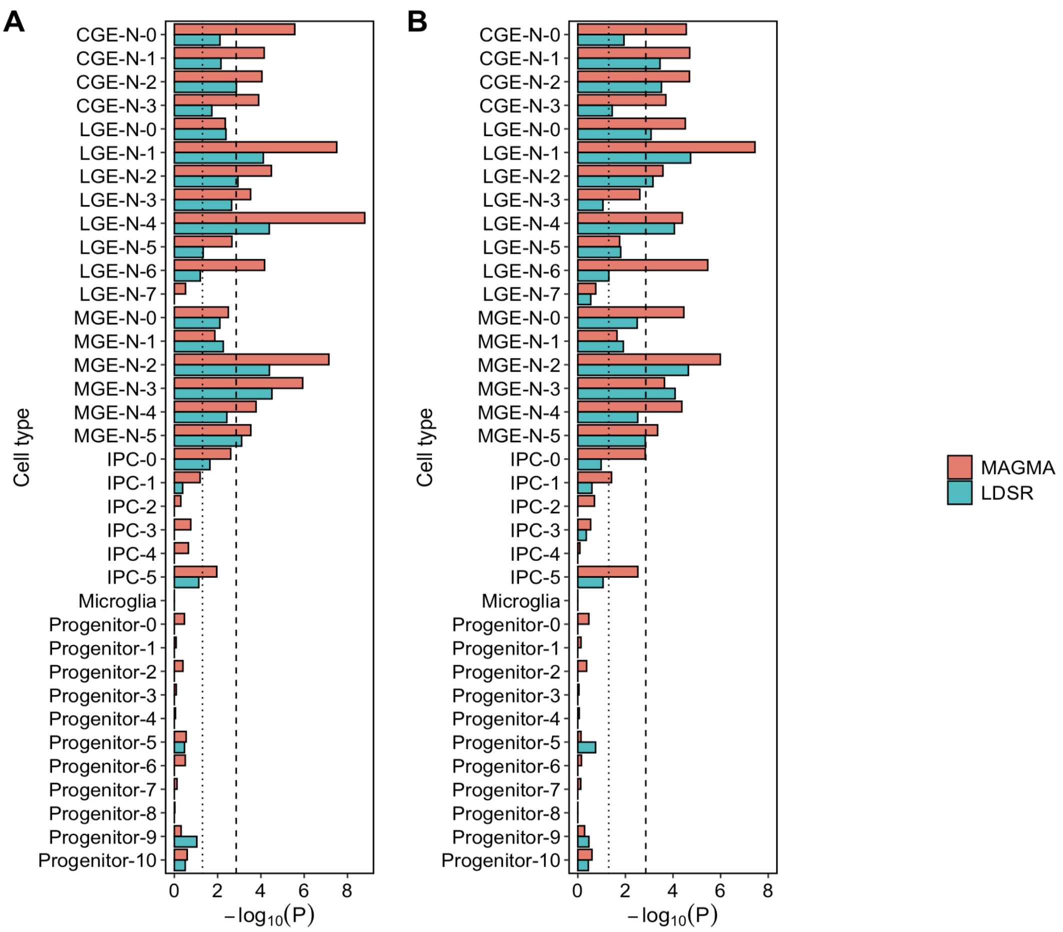


**Figure S7: Log10 *P*-values for enrichment of schizophrenia genetic associations in genes with high expression specificity for level 2 cell types of the ganglionic eminences using MAGMA and SLDSR after (A) downsampling the number of cells and (B) standardizing the number of genes included for each cell population.** A) -Log10 *P*-values when all level 1 cell types have been down-sampled to match the cell population with the lowest number of cells (microglia, with 242 cells). B) -Log10 *P*-values when the 1000 genes with the highest specificity scores for each cell population are analysed.The dotted vertical line indicates nominal ($P$ < 0.05) significance and the dashed vertical line indicates the Bonferroni-corrected *P*-value threshold for 36 tested cell populations at level 2 ($P < 1.4$ x$10^{-3}$). CGE-N = developing neurons from the CGE; LGE-N = developing neurons from the LGE; MGE-N = developing neurons from the MGE; IPC = intermediate progenitor cells.

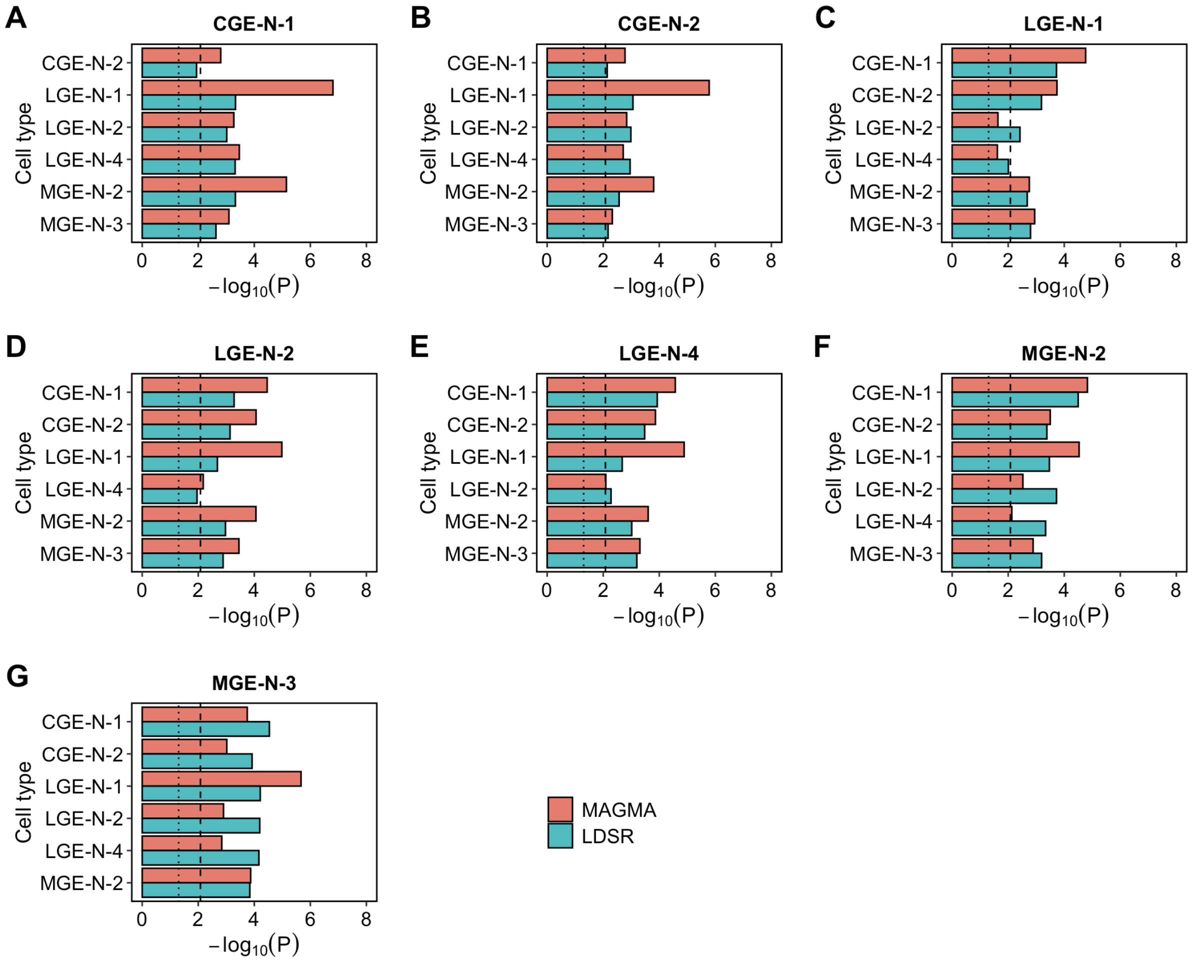


**Figure S8. -Log10 *P*-values for enrichment of schizophrenia associations in genes in the top expression specificity decile of each significantly enriched level 2 cell population of the ganglionic eminences, conditioning separately on genes in the top decile of each of the other 6 significantly enriched populations using MAGMA and LDSR**. The dotted vertical line indicates nominal ($P < 0.05$) significance and the dashed vertical line indicates the Bonferroni-corrected *P*-value threshold for 6 tested cell populations ($P < 0.0083$). CGE-N = developing neurons from the CGE; LGE-N = developing neurons from the LGE; MGE-N = developing neurons from the MGE.

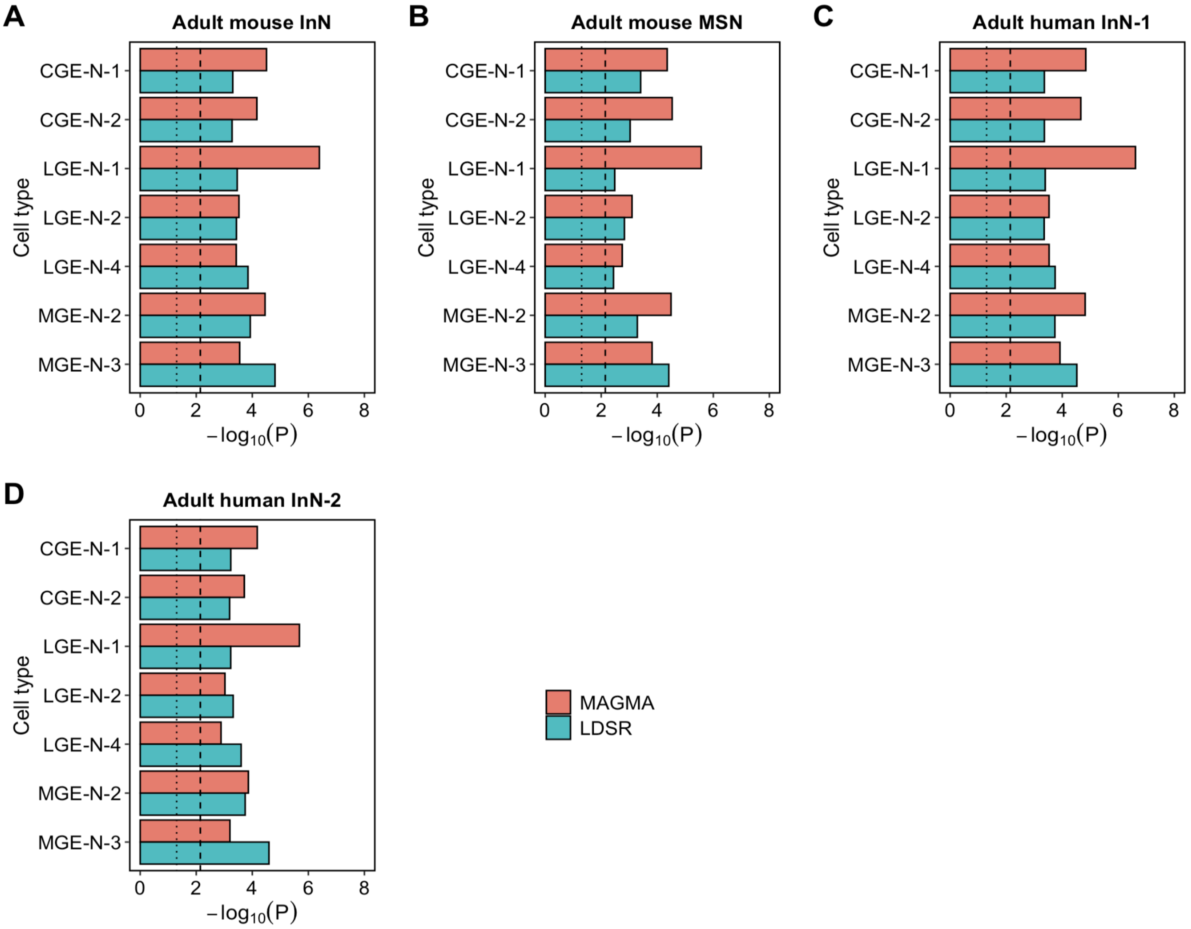


**Figure S9. -Log10 *P*-values for enrichment of schizophrenia associations in genes in the top expression specificity decile of significantly enriched level 2 fetal cell types of the ganglionic eminences, conditioning on genes in the top specificity decile of two schizophrenia-associated GABAergic neuron populations from adult mouse brain (mature interneurons of the cerebral cortex [InN] and medium spiny neurons of the striatum [MSN]) and two GABAergic neuron populations from the adult human brain (InN-1 and InN-2) (Trubetskoy et al, 2022) using MAGMA and SLDSR**. The dotted vertical line indicates nominal ($P < 0.05$) significance and the dashed vertical line indicates the Bonferroni-corrected *P*-value threshold for 7 tested cell populations ($P < 0.0071$). CGE-N = developing neurons from the CGE; LGE-N = developing neurons from the LGE; MGE-N = developing neurons from the MGE.

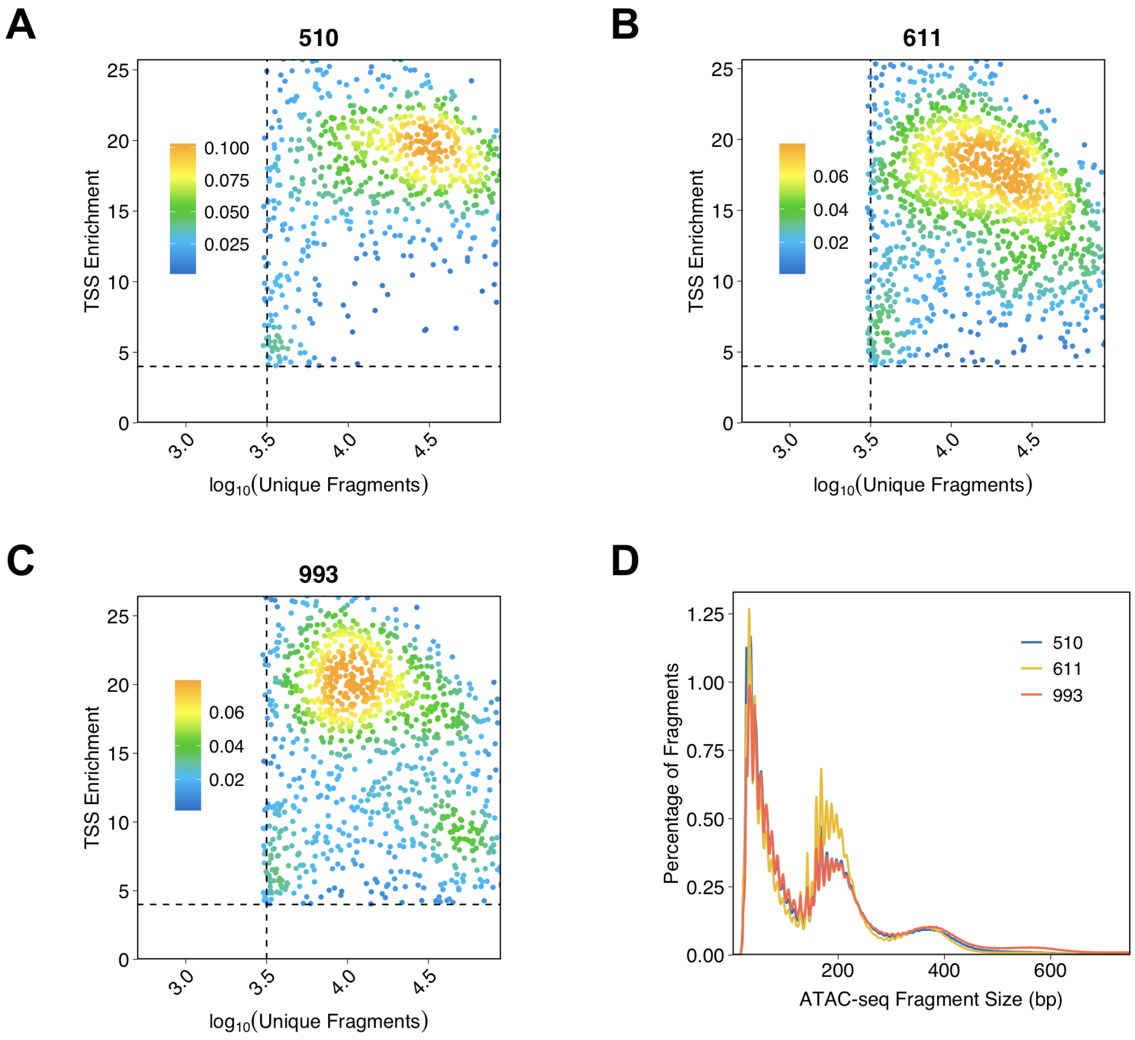


**Figure S10: Per sample quality control plots for snATAC-Seq data.** A-C) Transcription start site (TSS) enrichment and number of unique DNA fragments for snATAC-Seq data from nuclei retained from samples 510, 611 and 993. D) snATAC-Seq DNA fragment size from retained nuclei for each sample. Analyses performed using ArchR (Granja et al, 2021).

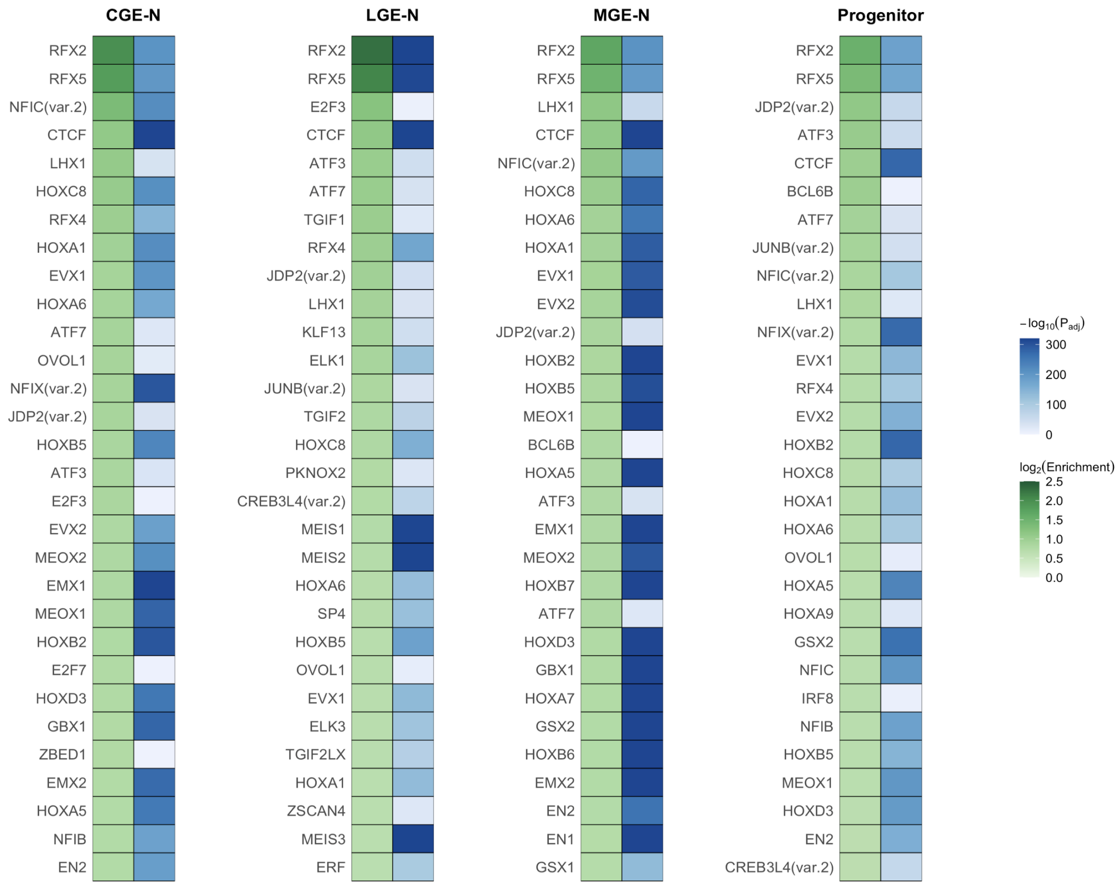


**Figure S11: The 30 most strongly enriched transcription factor binding motifs in open chromatin regions identified in each broad cell population of the ganglionic eminences.** Motifs are ordered by enrichment compared to randomly sampled, similarly sized sets of genomic sequences, accounting for differences in GC content and k-mer composition, using the R package monaLisa (Machlab et al, 2022). CGE-N = developing neurons from the CGE; LGE-N = developing neurons from the LGE; MGE-N = developing neurons from the MGE.

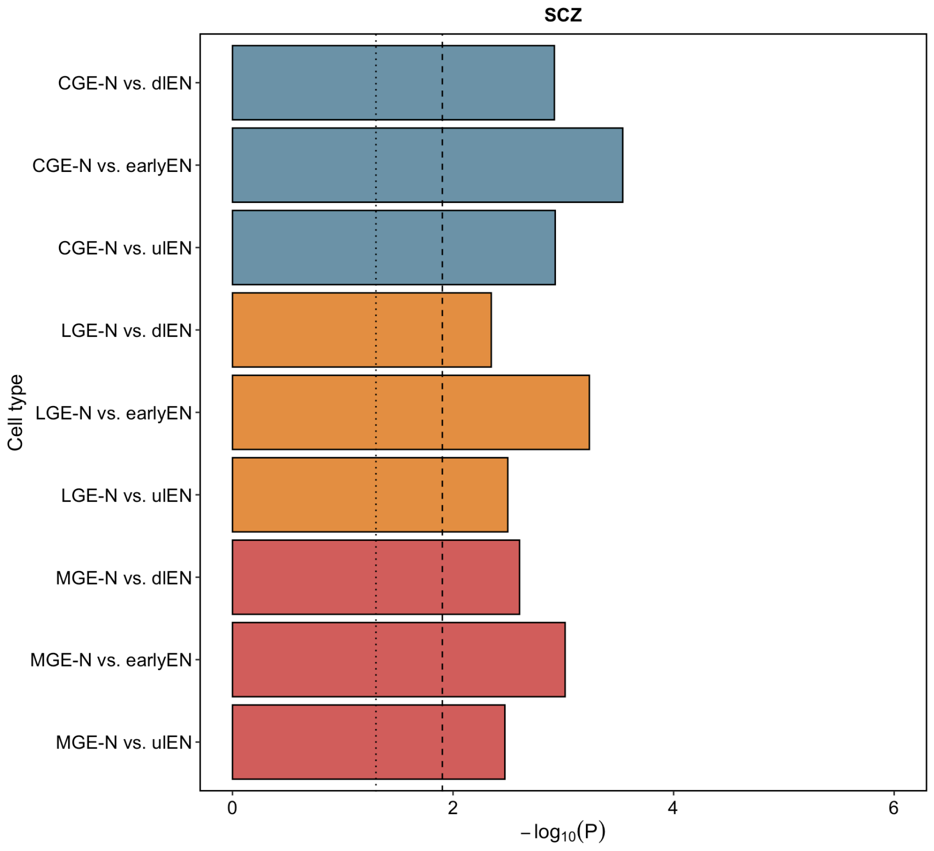


**Figure S12: -Log10 *P*-values for enrichment of schizophrenia SNP heritability in open chromatin regions of the ganglionic eminences, accounting for open chromatin regions identified in implicated excitatory neuron populations of the fetal brain (Ziffra et al, 2021), using SLDSR**. The dotted vertical line indicates nominal ($P < 0.05$) significance and the dashed vertical line indicates the Bonferroni-corrected *P*-value threshold for the 4 GE cell populations originally tested ($P < 0.0125$). CGE-N = caudal ganglionic eminence developing neurons; LGE-N = lateral ganglionic eminence developing neurons; MGE-N = medial ganglionic eminence developing neurons; dlEN = deep layer (cortical layers V–VI) excitatory neurons; earlyEN = early excitatory neurons; ulENs = upper layer (cortical layers II–IV) excitatory neurons.
